# Safety of administering biologics to IBD patients at an outpatient infusion center In New York City during the COVID-19 pandemic: Sars-CoV-2 seroprevalence and clinical and social characteristics

**DOI:** 10.1101/2021.03.15.21253615

**Authors:** Serre-Yu Wong, Stephanie Gold, Emma K. Accorsi, Tori L. Cowger, Dean Wiseman, Reema Navalurkar, Rebekah Dixon, Drew S. Helmus, CiTI Study Group, Adolfo Firpo-Betancourt, Damodara Rao Mendu, Susan Zolla-Pazner, Ken Cadwell, Jean-Frederic Colombel

## Abstract

Patients with immune-mediated inflammatory diseases (IMIDs) and acquired and genetic immunodeficiencies receiving therapeutic infusions are considered high risk for SARS-CoV-2 infection. However, the seroprevalance in this group and the safety of routine administrations at outpatient infusion centers are unknown. To determine the infection rate and clinical-social factors related to SARS-CoV-2 in asymptomatic patients with IMIDs and immunodeficiencies receiving routine non-cancer therapeutic infusions, we conducted a seroprevalence study at our outpatient infusion center. We report the first prospective SARS-CoV-2 sero-surveillance of 444 IBD/IMID, immunodeficiency, and immune competent patients at an outpatient infusion center in the U.S. showing lower seroprevalence in patients compared with the general population and provide clinical and social characteristics associated with seroprevalence in this group. These data suggest that patients can safely continue infusions at outpatient centers.

## Introduction

Due to their underlying conditions and frequent treatment with infusion medications that modulate the immune system, patients with inflammatory bowel disease (IBD) are considered high-risk for COVID-19. Since the beginning of the COVID-19 pandemic, clinical guidelines have advised patients to maximize shielding, and many patients receiving infusion therapies switched from infusion centers to home infusions or chose to delay or discontinue treatments.^1,2^ This places them at risk for treatment discontinuation, increased home exposures in the case of home infusions, and poor outcomes because of stopping therapies.^2,3^ However, the risk of SARS-CoV-2 infection in IBD patients receiving their care in an outpatient infusion center is unknown.

We thus screened patients for SARS-CoV-2 antibodies at our high-volume therapeutic infusion center in New York City during the height of the COVID-19 pandemic, including during lockdown when access to patients was limited, and assessed the contributions of social and clinical factors to seroprevalence. We reasoned that these patients might take greater shielding precautions due to COVID-19, which could contribute to seroprevalence, so we collected data on social exposures by a questionnaire.

## Methods

This was a cross-sectional study conducted from May 26 to July 15, 2020, at the Mount Sinai Therapeutic Infusion Center. We recruited adult patients aged 18 years and older (range 18-93) with scheduled appointments. All patients were negative for COVID-19 symptoms at the time of blood collection. For a general population control, we utilized previously-published SARS-CoV-2 serological testing data of 2,123 routine care patients at the same institution during the same time period. ^4^

Patient data including sex, infusion diagnoses, infusion medications, and lab values (hemoglobin, platelets, albumin, CRP, and ESR) were obtained from medical records. Participants were asked to complete questionnaires to report demographics, residential zip codes, medical history, COVID-19 symptom and testing history, and social exposures. Sera was tested using the (1) Emergency Use Authorization (EUA) Mount Sinai Health System Kantaro SeroKlir assay to compare results with data from the general population control, and (2) EUA Siemens Healthineers SARS-CoV-2 Total (COV2T) assay (Supplemental Methods).^4,5^ The Icahn School of Medicine at Mount Sinai Institutional Review Board approved the study. Statistical analysis was performed using R v3.5.3 (Supplemental Methods).

## Results

Overall 444 patients were recruited at the infusion center. The Kantaro SeroKlir results showed a 13.5% (52/383) SARS-CoV-2 seroprevalence in infusion center patients which was lower than 18.5% (393/2123) reported in a the general control population (p = 0.02).^4^

The Siemens COV2T assay, which has shown the highest sensitivity and specificity in comparative analyses, was used for all further analyses to broaden applicability of the data.^5^ Overall seroprevalence was 9% (40/444). Details of patient demographics, diagnostics and therapies are in Table 1. The seroprevalence was 9.4% (35/373) in IMID patients, including 9% in IBD (21/235), 22% (4/18) in immunocompetent patients, and 1.9% (1/53) in patients with acquired or genetic immunodeficiencies. Of IBD patients, seroprevalence was 7.8% in Crohn’s disease (11/141) and 10.6% in ulcerative colitis and IBD-unclassified (10/94) patients. Seropositive patients displayed the expected range of antibody titres (Figure 1).

**Table 1:**
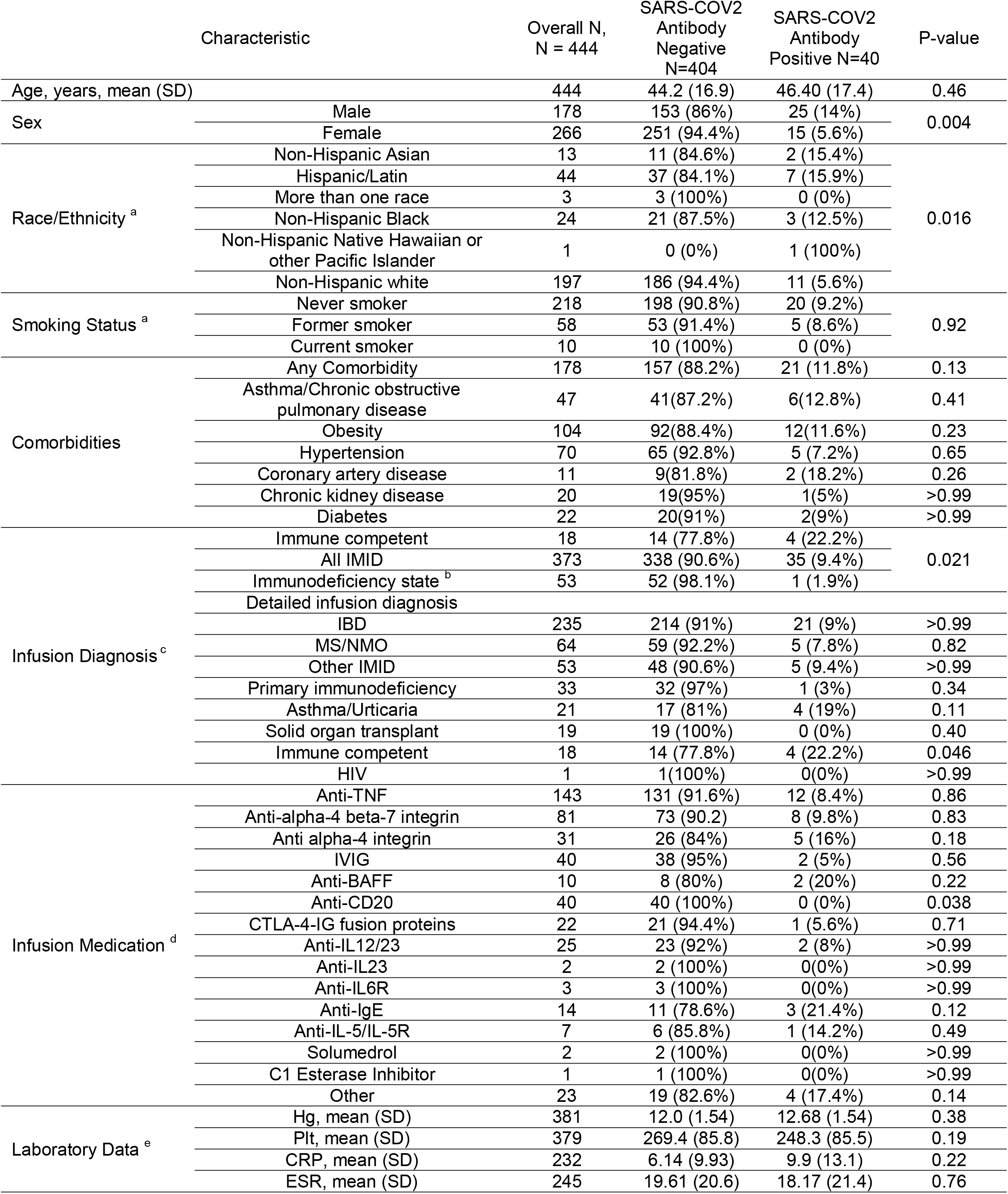
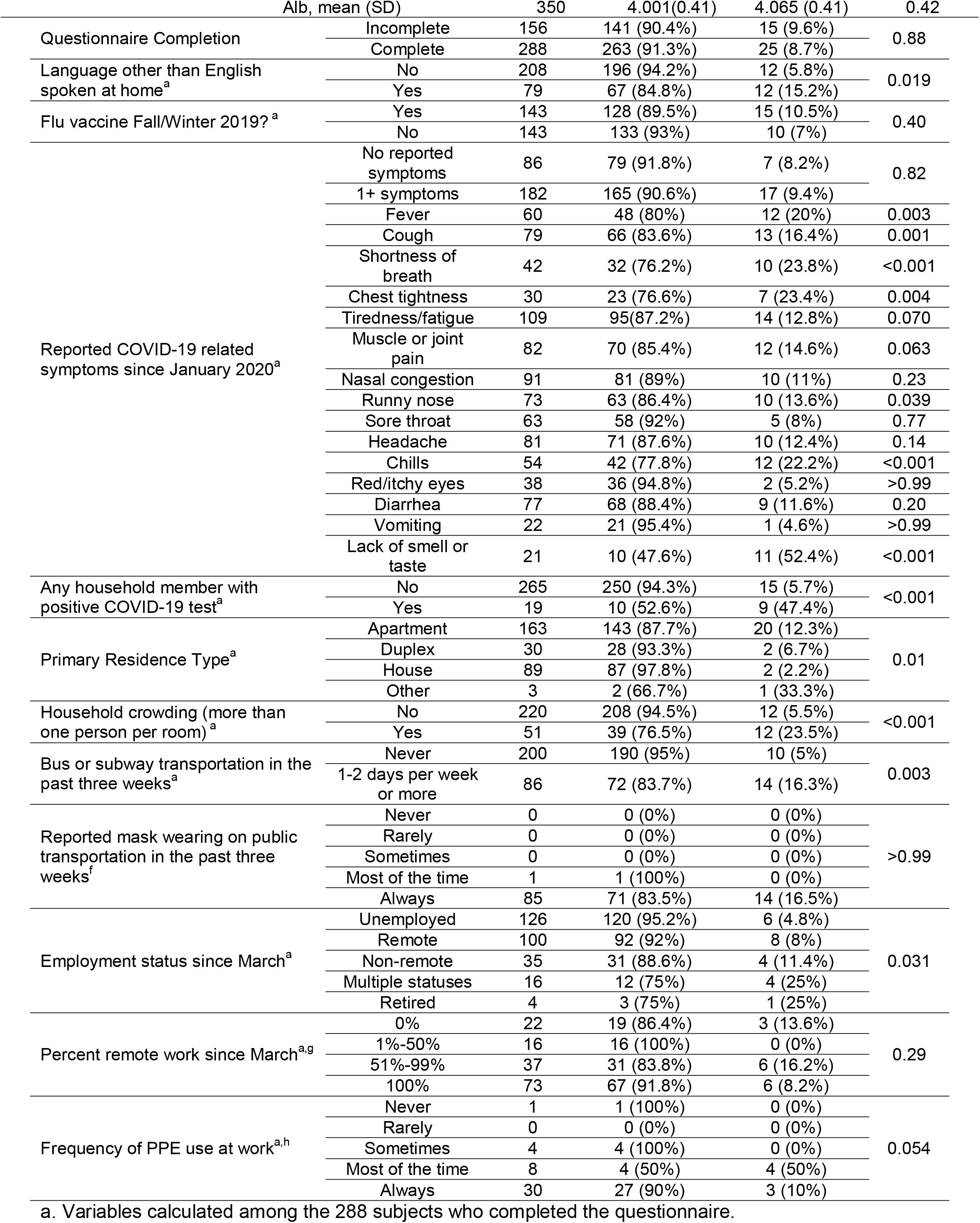
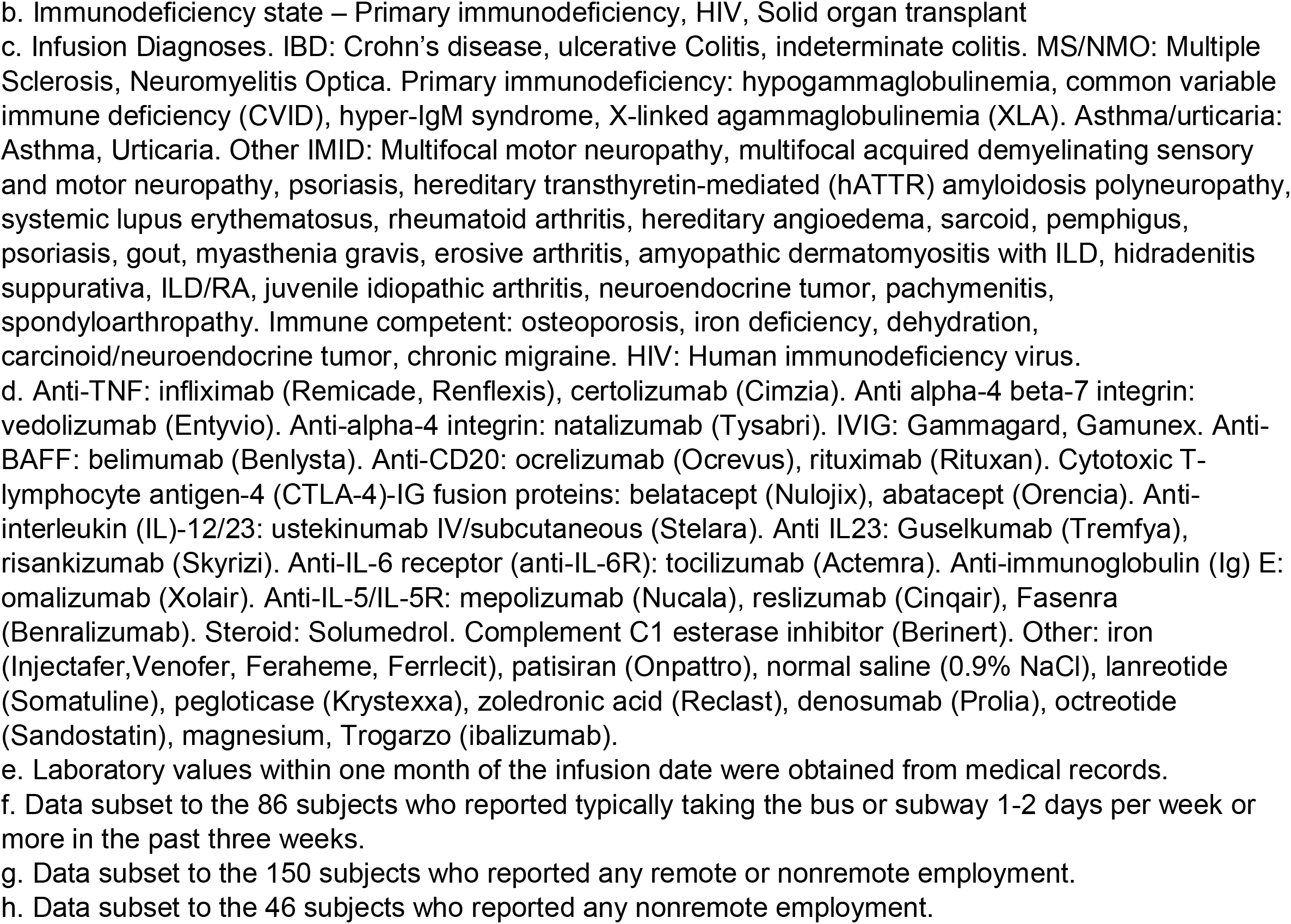
Selected Demographic Characteristics by SARS-CoV-2 Antibody Presence.

**Figure 1.**
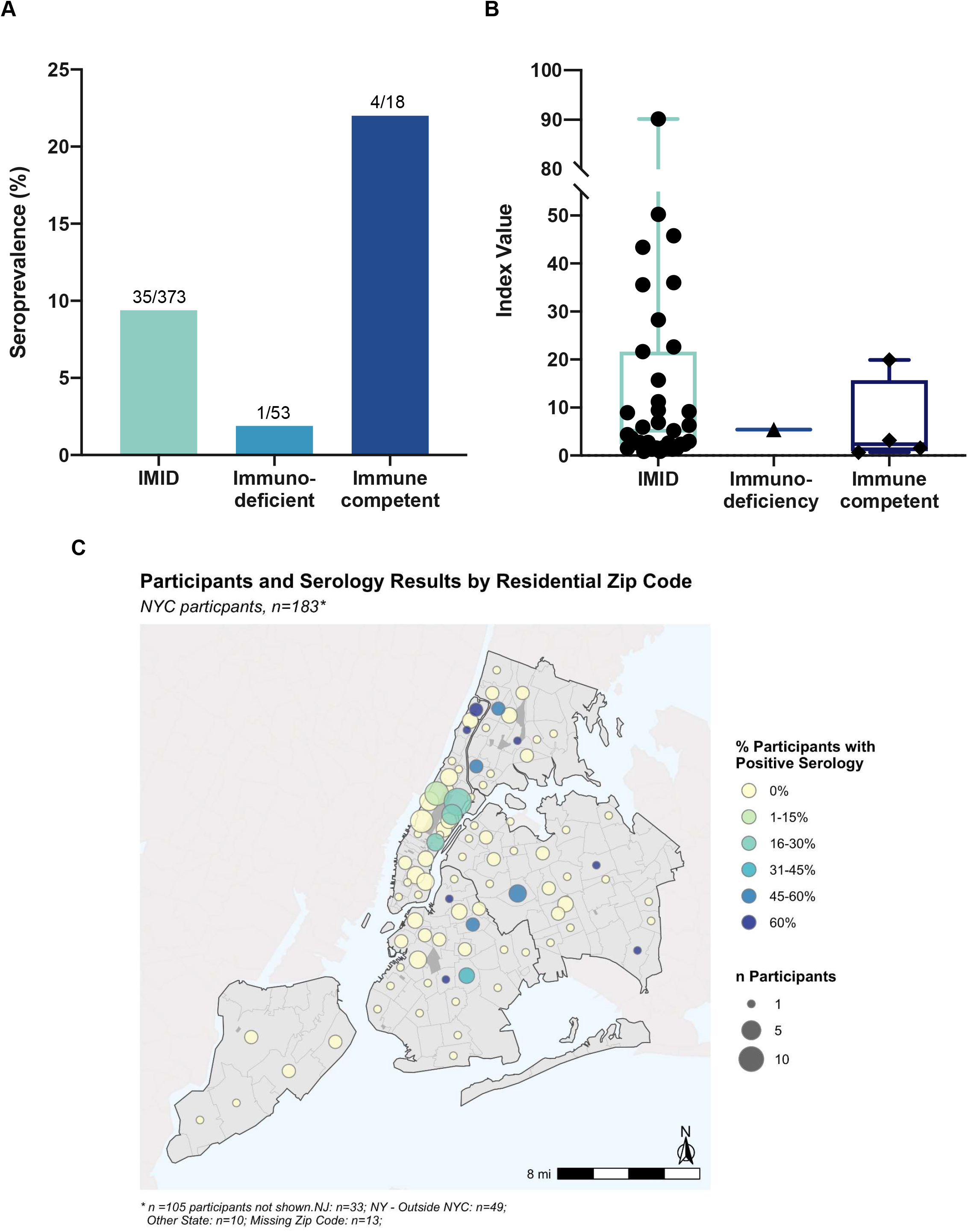
Seroprevalence of patients at the Mount Sinai Therapeutic Infusion Center. (A) Seroprevalence by diagnosis group. (B) Semi-quantitative IgG results for seropositive patients. (C) Seroprevalence by zip code of participants living in NYC.

The proportion of seropositive males (14%, 25/178) was higher than that of females (5.6%, 15/266). There were significant differences in seroprevalence by race and ethnicity, with the lowest seroprevalence in non-Hispanic white patients (5.6%, 11/197) compared with other groups. There were no significant associations with age, smoking status, or comorbid conditions previously established to be COVID-19 risk factors.

Among 288 of 444 patients who completed the questionnaire, SARS-CoV-2 seroprevalence was associated with COVID-19 symptoms and household member(s) testing positive for SARS-CoV-2 infection (PR: 8.37 [4.23,16.56], p < 10^−5^), speaking a language other than English at home (PR: 2.63 [1.23, 5.61], p = 0.019), household crowding (PR: 4.31 [2.06, 9.04], p < 10^−3^), primary residence type (p = 0.010), and use of public transportation (PR: 3.26 [1.51,7.04], p = 0.003). Of note, 25 of the 40 seropositive patients completed questionnaires, of whom 6 answered that they had no symptoms from January 2020 until the time of their blood draw, indicating asymptomatic seroconversion. Seroprevalence by reported home zip codes for 183 participants living in NYC was mapped (Figure 1). Mask wearing was 100% (43/43) among patients who reported non-remote employment and 98.5% (85/86) among those who utilized public transportation.

## Discussion

Our findings indicate that the SARS-CoV-2 seroprevalence of IBD patients receiving routine treatments at an outpatient infusion center is lower than that of a general population control. There was a high rate of mask wearing in this cohort, suggesting that shielding could have been protective. The analyses reveal that social, demographic, and geographic factors for SARS-CoV-2 infection that are well-characterized in the general population extend to IBD patients. To our knowledge, this is the first prospectively-collected seroprevalence data from a single large-volume non-oncologic outpatient infusion center at the height of the COVID-19 pandemic in New York City.

A study of German IMID patients receiving anti-cytokine therapies also showed a lower seroprevalence compared with control populations.^6^ Studies of IBD patients in Italy and Germany and the United Kingdom receiving infusion medications infliximab and vedolizumab showed that seroprevalence was not higher in IBD patients than controls.^7-9^ However, the general population seroprevalence in those regions were low.

Several important points can be made from the social questionnaire data. First, there was a high rate of mask wearing in this cohort, suggesting that shielding could have been protective. Second, these data substantiate the evidence for close, household contact as a primary risk factor for SARS-CoV-2 infection. Third, we confirm that well-characterized inequities by race/ethnicity and socioeconomic status extend to this cohort, and demonstrate that conditions related to work, housing, and transportation were also associated with increased prevalence of infection.^10^ These social and environmental conditions are shaped by structural racism and economic inequity and may explain observed inequities in COVID-19 burden. Fourth, analysis of zip codes shows higher seropositivity of patients living in areas of the New York City metropolitan area known to have higher rates of COVID-19 positivity, indicating seroprevalence in infusion center patients reflects the seroprevalence where they reside.

Strengths of this study include prospective data collection, first reporting of IBD receiving infusions at a large-volume outpatient center in the U.S., comparison of IBD patients to a general population control, other IMIDs, and immunodeficient patients, and comprehensive clinical and social exposure analysis. A limitation of the study is that it is from a single center.

In conclusion, our data suggest patients can safely continue routine infusions at outpatient centers during the COVID-19 pandemic as the seroprevalence at our center was low; longitudinal follow-up of these patients would confirm this point. This information is useful to practitioners as many patients have opted to delay or discontinue infusions during the pandemic, risking disease flares and complications thereof, or switched to home infusions, which has been associated with adverse outcomes.^2,3^ This is all the more important in our urban cohort given the disparity of seroprevalence by socio-economic factors. Finally, the functionality and durability of immune responses to SARS-CoV-2 infection, as well as COVID-19 vaccines, remain to be determined in IBD patients, who were not included in COVID-19 vaccine trials.

## Data Availability

This data will be shared upon request.

## Acknowledgments

The authors wish to thank the patients who participated in the study; the Mount Sinai Therapeutic Infusion Center staff for their contributions to patient recruitment; Siemens Healthineers and the Icahn School of Medicine at Mount Sinai Seronet Team for their support for serology testing; Gustavo Martinez-Delgado for laboratory support; and Rohit Chandwani, Manasi Agrawal, Viviana Simon, Ania Wajnberg, Saurabh Mehandru, Jack Satsangi, and Charlotte Cunningham-Rundles for thoughtful comments and discussions on study design and data analysis and presentation.

