## Supplementary Methods for "Safety of administering biologics to IBD patients at an outpatient infusion center In New York City during the COVID-19 pandemic: Sars-CoV-2 seroprevalence and clinical and social characteristics"

*Sample processing.* Blood specimens were collected in SST tubes, allowed to clot and centrifuged at 1100-1300*g* for 20 minutes at room temperature. The specimens were aliquoted into sterile cryovials and stored immediately at -80˚C until testing. Samples were transported on dry ice to Siemens Healthineers in Tarrytown, New York, where the COV2T assay was performed. The Siemens Healthineers SARS-CoV-2 Total (COV2T) assay is a chemiluminescence-based assay, which measures total antibodies to SARS-CoV-2 RBD. Positive samples were additionally tested by a semi-quantitative assay for anti-RBD immunoglobulin (Ig)G. The Kantaro SeroKlir assay was established by the Mount Sinai Health System CLIA-certified Clinical Pathology Laboratory, which is a two-step ELISA protocol that first screens for antibodies to the SARS-CoV-2 Spike protein RBD followed by a confirmatory ELISA for presumed positives measuring antibody presence and titres for full-length, recombinant Spike protein.

*Statistical methods.* To identify associations between antibody presence and clinical, demographic, and social covariates, risk ratios and 95% confidence intervals were calculated using unconditional maximum likelihood estimation (Wald) and normal approximations (Wald), respectively, in the “epitools” package. For dichotomous and categorical covariates, p-values were calculated using Fisher’s exact test when the expected frequency of one or more cells was less than 5, and the chi-square test with Yates continuity correction otherwise. For continuous covariates, p-values were calculated using Student’s t-test. Harvard T.H. Chan School of Public Health Institutional Review Board approved secondary data analysis for questionnaire data.

*Additional questionnaire methods.* For household crowding, participants were asked “How many people live in your household, excluding you?” and “How many rooms are there in your primary residence?” Household crowding was defined as having more than one household member per room with the number of people per room calculated as: (1 + number of other household members) / number of rooms. For public transportation usage, participants were asked “In the past three weeks, how many days did you typically take the bus?” and “In the past three weeks, how many days did you typically take the subway/short-distance train?” Participants were considered to have ridden public transportation if they selected any answer other than “never” to either question. For race/ethnicity, participants were coded as Latinx if they reported Hispanic ethnicity and any race. All participants who did not report Hispanic ethnicity were then coded by race. The frequency of mask usage was measured on a 5-point scale consisting of “never”, “rarely”, “sometimes”, “most of the time”, and “always”.
